# Recovery from the COVID-19 pandemic by mass vaccination: emergent lessons from the United States and India

**DOI:** 10.1101/2021.05.26.21257847

**Authors:** Edwin Michael, Ken Newcomb, Anuj Mubayi, Imran Mahmood

## Abstract

The advent of vaccinations has heightened global optimism that the end of the SARS-CoV-2 pandemic could be in sight. However, concerns, including the impact of variations in the rates of vaccination between countries, raise questions about the use of mass vaccination for accomplishing a quick recovery from the contagion. Here, we used a SEIR-based model calibrated to data on the pandemic and vaccinations reported for the United States (US) and India to gain strategic insights into using mass vaccinations for ending COVID-19. We estimate that while up to 65% of the US population is already immune to the virus due to the recent rapid mass vaccinations carried out, only 13% of the Indian population may be immune currently owing to a slow rate of vaccination and the effect of a stricter lockdown imposed to curb the first wave of the pandemic. We project that due to the higher immune to susceptible ratio already achieved in the US, the pandemic will only decline if the present rates of vaccinations and social mitigations are continued and remain effective. By contrast, the recent loosening of social measures coupled with a slow rate of vaccination is the chief reason for the virus resurgence in India, with only immediate lockdowns coupled with ramping up of vaccinations providing the means to control the present wave. These results highlight that using mass vaccination to achieve a speedy recovery from the SARS-CoV-2 pandemic will depend crucially on the ability to carry out national vaccinations as rapidly as possible.

## Main Text

### Introduction

As vaccinations against severe acute respiratory syndrome coronavirus 2 (SARS-CoV-2) roll out in countries (1), interest is growing in how best to use the current vaccines for rapidly ending the COVID-19 pandemic (2). Indeed, with cases and hospitalizations beginning to fall in countries that appear to have vaccinated the largest shares of their populations, there is optimism that vaccinations may portend the endgame for the pandemic (3). However, vaccine coverages to protect against infection above 80% are thought to be desirable (4), and this, coupled with variation in the rates of vaccination achieved between countries (1), supply-chain and distributional challenges, uncertainties regarding the protective efficacy of individual vaccines against different virus variants (5), and the durability of the induced immunity (2,4), continue to raise questions about the use of mass vaccination for affecting a quick recovery to normalcy from COVID-19 (2).

Empirical evaluations of associations between vaccination rates and reported daily new COVID-19 cases indicate high variability between countries, although there is some direct evidence that the group of countries that have achieved the highest vaccination rates to date globally also have the lowest reported new cases, and consequently hospitalizations and deaths, post initiation of vaccination programs (3). This lack of a clear pattern from empirical data suggests that many other factors are likely to confound the relationship between mass vaccinations and infection incidence, including histories of social mitigation measures, spread of new more contagious variants, and prevailing levels of population immunity (4-6).

Here, we use a Monte Carlo-based SEIR-type COVID-19 mathematical model, incorporating the dynamics of imperfect vaccines and social mitigations (7), sequentially calibrated to data on historical and recent changes in cases, deaths and vaccination rates, reported for the United States (US) and India - two large countries with contrasting socio- economic conditions that are also at the extremes of the current state of the pandemic and vaccination effort - to gain strategic insights into the prospects as well as challenges to using mass vaccination as a tool for ending SARS-Cov-2 infection safely and rapidly.

## Results and Discussion

Fig. 1 depicts the vaccination rates, fractions of the population presently immune and the times at which herd immunity (if immunity is assumed reasonably long-term (10)) can be expected to be accomplished with the prevailing vaccines in both the US and India. These results indicate that while the proportion of the population vaccinated in the US has already reached 45% - as a result of the recent rapid mass vaccinations carried out in that country - only about 9% of the corresponding Indian population have been vaccinated to date. However, inclusive of acquired immunity, we also predict that 65% of the US population may already be immune to the virus compared to just 13% of the Indian population. This difference in the population immunity levels achieved between the two counties undoubtedly reflects the faster rate with which vaccinations were carried out in the US compared to India; however it also indicates the effect that differing levels of social mitigation measures imposed in the two countries during the course of their respective pandemics have had on the evolution of population immunity (Fig. 1). Thus, while India imposed a draconian lockdown to successfully curb the first wave of the pandemic (11), a significant negative epidemiological trade-off of this approach has been that this has resulted in a critically low rate of development of natural immunity to the virus by the end of the first wave compared to the US where earlier lockdowns were comparatively less stringent (compare Fig. 1*A* and *B*). This means that while the US has reached a stage where the fraction of the population immune now is greater than the fraction susceptible, a large susceptible pool was left intact in India due to both the strict lockdown imposed and a slow vaccination rate, leaving the country highly vulnerable to virus resurgence as a result of any loosening of the imposed social mitigation measures.

**Figure 1.**
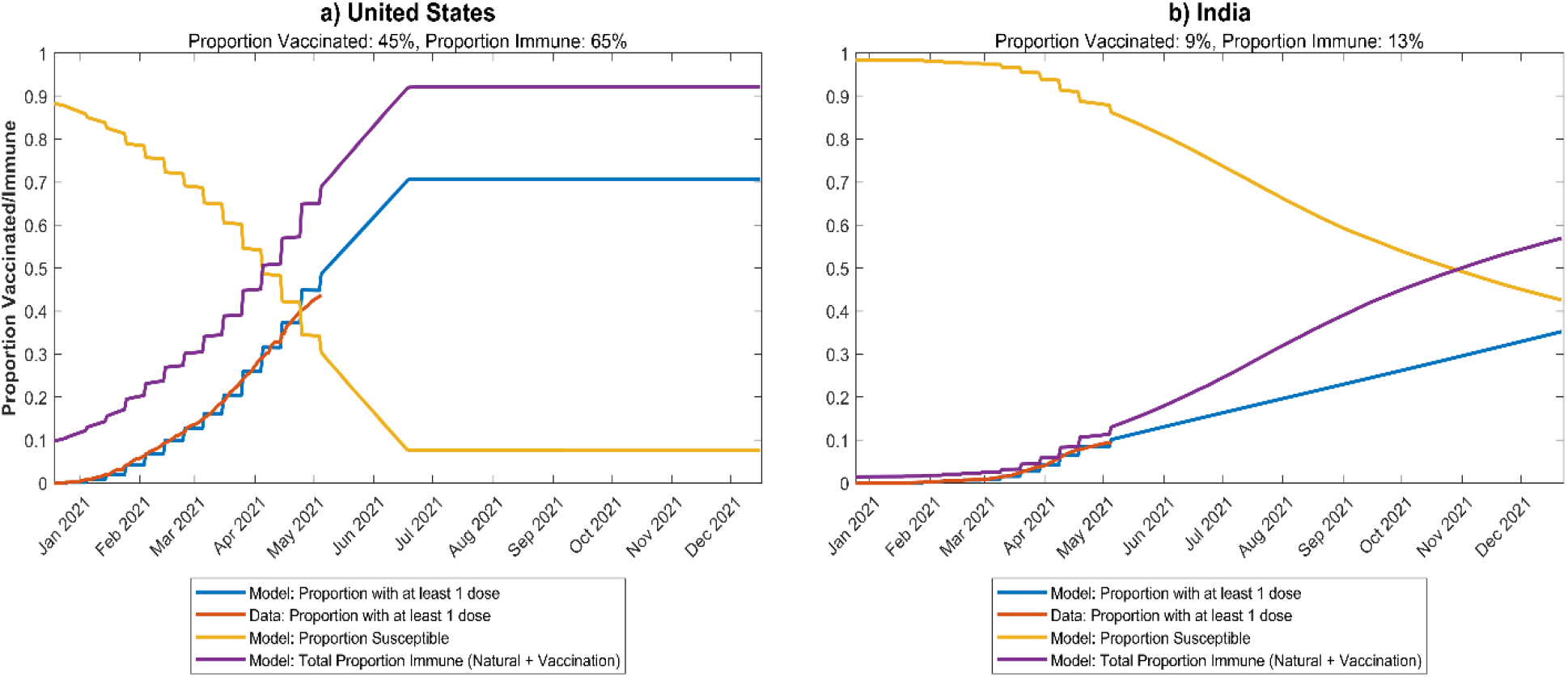
Model predictions of the proportions vaccinated, susceptible, and immune plotted alongside reported vaccination data for the United States (a) and India (b). Model predictions for five scenarios are shown: a) continuation with current social measures and vaccination rates (median along with 90% confidence levels), b) a 30-day 25% increase in social measures alongside current vaccination rates, c) a 30-day 50% increase in social measures alongside current vaccination rates, d) a 30-day 25% increase in social measures, with a weekly 50% ramp up of vaccinations up to 3x the current rate, and e) a 30-day 50% increase in social measures, with a weekly 50% ramp up of vaccinations to 3x the current rate. The proportion immune in the United States due to vaccinations at this time is predicted to be 45%, while inclusive of naturally acquired immunity, the overall proportion immune could be as high as 65%. The respective proportions for India are estimated to be only 9% and 13%.

Indeed, our projections in Fig. 2 support the above conclusions amply. These show that as a result of the higher immune to susceptible ratio achieved in the US, the pandemic will only decline and fade out going forward if current vaccines remain effective and present vaccination rates and social measures are continued. By contrast, the recent loosening of social measures that began in March 2021 coupled with a low vaccination rate (12) is the chief reason for the alarming resurgence of the virus in India. Our results show that only by the imposition of an immediate short duration lockdown (month-long) in the affected states, as has been done by several affected states, will India be able to control the current wave, while ramping up the current vaccination rate alongside this lockdown - in a 2-week stepped fashion until approximately the 3.15 million daily vaccination rate that was conducted in April 2021 is achieved by end of the lockdown - will also allow the country to end the pandemic by the end of the year. Continuing with the current vaccination rate in this scenario will by contrast lengthen the time to end the pandemic (Fig. 2*B*). Note also, that if vaccinations are not ramped up alongside the proposed lockdown, third albeit smaller but still sizeable resurgent third waves will occur in the country during the coming fall, which might necessitate the imposition of further economically-and-socially damaging lockdowns (Fig. 2*B*).

**Figure 2.**
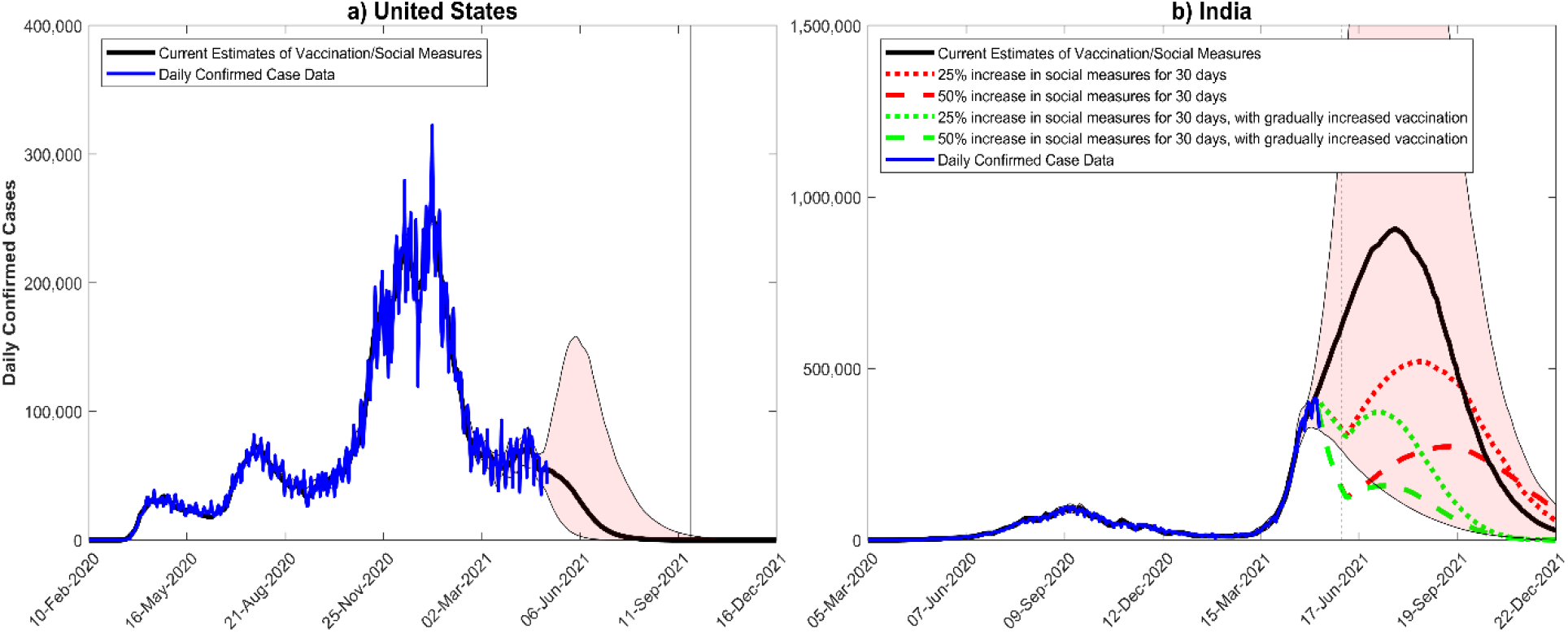
Projections of the daily confirmed cases for the United States (a) and India (b) for different vaccination and social mitigation scenarios. The blue curves denote reported case data while the solid and dotted or dashed curves present model forecasts for the various scenarios described in the respective legend boxes. For the US, the median daily confirmed cases is projected to drop below a single case on September 18^th^, 2021 if current vaccination rate/social measures are continued and remains effective against the virus. By contrast, median cases in India are forecasted to increase exponentially and peak on July 22^nd^ with 900,000 confirmed cases in the absence of any increase in social distancing measures or vaccination (black curve), although peak daily cases could approach > 2 million going by the upper 90% confidence interval predictions (grey band). Imposing an immediate short-term lockdown for a month will reduce incidence quickly, but significant third waves during the fall would occur if this is unaccompanied by a ramp up of the current vaccination rate (∼1.1 million daily doses). Increasing the current vaccination rate 3x in this scenario (by increasing the rate in 2-week increments to 1.5x, 2x, 2.5x and 3x the current rate) would prevent these waves and also cause the pandemic to end by year end.

Overall, these results highlight the importance of achieving the rapid vaccination of the population, as accomplished in the US and other countries, such as Israel and the UK (1,3) as key to using mass vaccinations for accomplishing a quick recovery from the pandemic while current vaccines are still effective against prevailing virus variants (5). By contrast, the lesson from India is that it will be too dangerous to reopen the economy while vaccination rates are still low, transmission is ongoing, and a large fraction of the population remains susceptible to infection. These results suggest that we need to focus global and national attention on increasing the production, procurement, and delivery of the existing COVID-19 vaccines expeditiously to all affected populations if we are to effectively recover from the present pandemic as speedily as possible.

## Materials and Methods

We extended our data-driven SEIR-based COVID-19 model (7) to include the dynamics of imperfect vaccines and impacts of social mitigation measures to perform the present simulations. Briefly, the model simulates the course of the pandemic in each country via the adaptive rate of movement of individuals through various discreet compartments, including different infection and symptomatic categories as well as immune, vaccination and death classes (7). We also assume that each population is closed and that their population sizes remain constant over the duration of the simulations reported here. Calibration of the model to capture the transmission conditions of each country was performed by fitting the SEIR model sequentially to daily confirmed case, death, and vaccination data assembled from the start of the epidemic until May 5^th^ 2021, as provided by the The Coronavirus App (https://coronavirus.app). A 7-day moving average is applied to the daily confirmed case and death data to smooth out fluctuations due to COVID-19 reporting inconsistencies.

A sequential Monte Carlo-based approach was used for carrying out the updating of the model by first sampling 50,000 initial parameter vectors from prior distributions assigned to the values of each parameter for every 10-day block of data (7). An ensemble of 250 best-fitting parameter vectors, based on a Normalized Root Mean Square Error (NRMSE) (8) between predicted and observed case and death data, is then selected for describing these 10-day segments of data. Updating of the parameters is then accomplished by using the best-fitting ensemble of parameter posteriors as priors for the next 10-day block, and the fitting process is repeated. In addition, 50% of parameters is drawn from the initial prior distribution to avoid parameter depletion during each updating episode (9). The strength of social distancing measures imposed by authorities to limit contacts is captured through the estimation of a scaling factor, *d*, which is in turn multiplied by the transmission rate, *beta*, to obtain the population-level transmission intensity operational at any given time in a population (7). This factor accounts for the effects of mask wearing, reductions in mobility and mixing, work from home, and any other deviations from the normal social behavior of a population prior to the epidemic.

The vaccination data for the US and India are directly applied by moving the proportion of the population that is vaccinated over a 10-day time interval from the susceptible class to the vaccine (1^st^ dose) class. Individuals then move from the vaccine to the booster (2^nd^ dose booster) class at a daily rate approximating a 21 day interval between vaccine doses. We assume a 1^st^ dose efficacy against acquisition of infection (the degree protection) to be 75% while this is raised to be 90% following the 2^nd^ booster dose. For the US simulations, we further consider that a fraction (conservatively set at 10%) of the susceptible population will refuse vaccinations, and we simply move this fraction to a new susceptible but vaccine refusing class that otherwise behave similarly as the main susceptible class. Average vaccination rates estimated from the last 3 weeks of the vaccination data in each country (April 15^th^ -May 6^th^) were used to simulate into the future. The equations governing the evolution of the system, their prior and posterior fitted parameter values, and the model code used to perform the simulations are available at: https://github.com/EdwinMichaelLab/COVID-SEIR-India.

## Data Availability

The computation system, data and model parameters are available at: https://github.com/EdwinMichaelLab/COVID-SEIR-India

